# Identification of a Transcriptional Hippo Response Signature Connecting Inflammatory Bowel Disease and Colorectal Cancer Progression

**DOI:** 10.64898/2026.09.24.26363887

**Authors:** K Sai Vignaish, Pranali Joshi, Sana Noor

## Abstract

**Background:** Inflammatory bowel disease (IBD) has been linked to a two- to eightfold increase in colorectal cancer (CRC) risk. However, particular genes that bridge intestinal dysbiosis and oncogenic transcriptional reprogramming remain unclear. The Hippo/YAP pathway is a major regulator of intestinal epithelial homeostasis whose effector YAP is hyperactivated in CRC, but its transcriptional engagement in IBD mucosa and its link to specific cancer-risk genes has not been thoroughly mapped.

**Methods:** Bulk RNA-seq data were integrated from two independent cohorts: GSE235236 (IBD colon mucosa; UC/CD vs. healthy controls; n = 56) and TCGA-COAD/READ (CRC tumors stratified by YAP activity; n = 592). Differential expression analysis was conducted separately within each cohort using Welch’s t-test with Benjamini-Hochberg FDR correction. YAP transcriptional activity was estimated per sample as the mean row-wise Z-score of seven canonical YAP target genes, and TCGA tumors were stratified into Hippo-High (Q4) and Hippo-Low (Q1) groups. Genes showing consistent dysregulation across both cohorts were identified and ranked to define a Dysbiosis-Hippo Response (DHR) module, which was further organized into functional gene archetypes.

**Results:** IBD mucosa showed 2,848 significantly dysregulated genes (FDR < 0.05). Comparison with the CRC cohort identified 120 genes showing concordant dysregulation, forming the DHR module. These genes were significantly enriched in TNF-alpha signaling via NF-kB (adjusted p = 0.006), inflammatory response (adjusted p = 0.013), and Myogenesis (adjusted p = 0.013). DHR gene classification revealed three functional archetypes: (i) innate immune activation (SPI1, OSM, IL11, LILRA2, SLC11A1) reflecting convergent myeloid reprogramming; (ii) epithelial-mesenchymal transition (TWIST1, SNAI1, PRRX2, MMP9) priming invasive potential before malignancy; and (iii) tumor microenvironment immune regulation (TN-FRSF4, TNFRSF18, PDCD1, VASN, SPON2) shaping immune tolerance. Interestingly, the canonical YAP target genes (CTGF, CYR61, ANKRD1, AMOTL2, and FSTL1), sharply upregulated in Hippo-High CRC, were essentially unchanged in IBD mucosa. This signifies that the shared DHR programme operates through non-canonical YAP effectors. And yet, YAP target scores are positively correlated with DHR module expression across IBD samples (Spearman rho = 0.726, p = 2.5 x 10-10).

**Conclusions:** Our findings define a three-archetype DHR module that links gut dysbiosis to YAP-driven oncogenesis at single-gene resolution. The programme shared between IBD and CRC is driven by innate immune activation, EMT priming, and remodeling of the immune microenvironment, not by the canonical CTGF/CYR61 YAP target axis, which appears only at the CRC stage. This gene-level framework provides tractable mechanistic targets for chemoprevention and biomarker development at the inflammation-to-cancer transition.

## Introduction

Colorectal cancer (CRC) is the third most common cancer worldwide and a major cause of mortality. Individuals diagnosed with inflammatory bowel disease (IBD), which incorporates ulcerative colitis (UC) and Crohn’s disease (CD), face a two- to eightfold elevated lifetime CRC risk relative to the general population, with risk proportional to disease duration and the more their bowel is affected [1, 2]. The progression from chronic mucosal inflammation to dysplasia and eventually to carcinoma is driven by constant damage to the gut lining and a weakened intestinal barrier. In addition, DNA-damaging molecules (reactive oxygen and nitrogen species) released during active inflammatory flares drive this transition [3].

One hallmark of IBD is gut dysbiosis, an imbalance of the colonic microbiome characterized by depletion of butyrate-producing bacteria (such as Faecalibacterium prausnitzii), concurrent with overgrowth of pro-inflammatory species and disruption of colonocyte metabolic homeostasis [4]. Although observational and experimental data have established a strong connection between this dysbiosis and increased CRC risk, the specific genes that modify microbial signals and reprogram colonic epithelial cells to an oncogenic transcriptional state are poorly defined.

The Hippo signaling pathway is a highly conserved kinase cascade that plays a central role in controlling organ size, tissue homeostasis, and epithelial regeneration [5]. When signalling is active, the kinase cascade MST1/2 –> LATS1/2 –> YAP/TAZ phosphorylates the transcriptional co-activators YAP (encoded by YAP1) and TAZ (encoded by WWTR1), keeping them sequestered in the cytoplasm and making them for proteasomal degradation.

When the cascade is suppressed, unphosphorylated YAP and TAZ translocate to the nucleus, where they partner with TEAD family transcription factors (TEAD1-4) to switch on canonical target gene programmes, including CTGF (CCN2), CYR61 (CCN1), ANKRD1, AREG, BIRC5, AMOTL2, and FSTL1-together driving cell proliferation, ECM remodeling, immune evasion, and invasion [6]. In colorectal cancer, YAP is hyperactivated in the majority of tumors, and its overexpression correlates with lymph node metastasis, advanced TNM stage, and poorer overall survival [7, 8].

There are various routes through which converging signals in the IBD microenvironment awaken YAP-bypassing the canonical Hippo cascade entirely. One of the first casualties is short-chain fatty acid signaling; when butyrate-producing commensals are depleted, it removes a key suppressor of YAP nuclear localization in colonocytes [10]. On top of that, the pro-inflammatory cytokines elevated in IBD mucosa activate YAP through NF-kB pathway crosstalk and cytoskeletal tension [11]. Mucosal oedema during IBD flares adds mechanical strain on colonocytes, activating YAP through integrin and Rho-GTPase mechanosensing [13]. At the individual gene level, the acetaldehyde produced by dysbiotic bacteria (in the absence of ADH1C-mediated detoxification) induces DNA damage responses known to suppress LATS1/2, creating a molecular bridge from microbial dysbiosis to YAP target gene induction.

Earlier transcriptome studies concerning IBD and CRC have mostly been conducted independently. The key unresolved question is which specific genes simultaneously carry dysbiosis-driven transcriptional imprinting in IBD mucosa and drive oncogenesis in established CRC. We therefore developed an integrative framework that (i) identifies genes concordantly dysregulated across IBD and Hippo-active CRC, (ii) ranks them into a Dysbiosis-Hippo Response (DHR) module, and (iii) classifies each gene into functional archetypes that translate statistical overlap into mechanistic hypotheses. We further examine whether YAP pathway activity in IBD quantitatively predicts DHR module expression.

## Methodology

### Datasets and Data Access

IBD cohort (GSE235236): GEO (accession GSE235236) provided the colon mucosal RNA-Seq data. RSEM-normalized expression profiles from colonoscopic biopsies of patients with UC (n = 26), CD (n = 22), and healthy controls (HC; n = 8; total n = 56) are included in the dataset. No further ethical approval was needed for reanalysis because all samples were de-identified. TCGA colorectal cancer cohort: cBioPortal (https://www.cbioportal.org) provided Bulk RNA-seq expression data (RSEM) and clinical metadata for TCGA-COAD and TCGA-READ. As the TCGA-CRC cohort comprises tumor samples without matched adjacent normals, all analyses were performed within the tumor space using YAP-based stratification as the activity contrast. After quality filtering, 592 were left.

### Differential Gene Expression Analysis of IBD

The log2-transformation was applied to the raw expression matrix as log2(TPM + 1). Patients with UC and CD were merged into one IBD group (n = 48). Welch’s test with BH-FDR correction was used to evaluate differential gene expression between IBD and HC. Significance threshold: FDR < 0.05 and |log2FC| >= 0.5. Results stored in IBD_DEG_IBD_vs_Control.csv.

### TCGA YAP Activity Scoring and Tumour Stratification

The log2-transformed TCGA RSEM values were expressed as log2(RSEM + 1). The mean row wise Z-score for each of the seven canonical YAP/TEAD target genes-CTGF, CYR61, ANKRD1, AREG, AMOTL2, BIRC5, and FSTL1 was used to calculate a per-sample YAP target score.

According to the CRC transcriptomics convention, “Hippo-High” indicates tumors with high YAP/TAZ target gene output (Q4 YAP target score), indicating functionally active YAP downstream signaling. Tumors were stratified into Hippo-High (Q4, n = 148) and Hippo-Low (Q1, n = 148); the middle 50% were excluded in order to maximize expression contrast. DEG between groups: Welch’s t-test/BH-FDR. Results stored in TCGA_DEG_HippoHigh_vs_HippoLow.csv.

### Cross-Cohort Overlap and DHR Module Construction

Significant genes from each cohort (FDR < 0.05, |log₂FC| >= 0.5) were inner-joined using the gene symbol. The sign of log₂FC was used to measure direction concordance; concordant genes were carried forward. A combined significance score was computed:

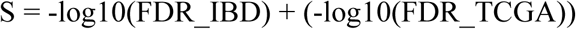

Simultaneous statistical robustness in both cohorts is indicated by higher S. The DHR module consists of the ranking concordant gene set. Results stored in core_signature_genes.csv.

### Classification of DHR Gene Functional Archetypes

Each DHR gene was annotated based solely on genes confirmed within the 120-gene concordant overlap. Annotations were based on (i) known function in mucosal immunity or intestinal homeostasis; (ii) established role in CRC or the YAP/Hippo transcriptional pathway; and (iii) relevance to gut microbiome biology and tumour microenvironment. Genes were categorized into three functional archetypes: Innate Immune Activation, Epithelial–Mesenchymal Transition (EMT) and Invasive Priming, and Tumour Microenvironment Immune Regulation. In both cohorts, all archetype gene annotations are restricted to genes with FDR < 0.05 and |log2FC| ≥ 0.5.

### Functional Enrichment Analysis

Enrichment of the DHR module was assessed using gseapy / Enrichr API across GO Biological Process 2021, KEGG 2021 Human, and MSigDB Hallmark 2020. Threshold: adjusted p < 0.05. Results stored in core_signature_enrichment.csv.

### Analysis of Mechanistic Association in IBD Samples

Using identified YAP target genes (ANKRD1, AREG, AMOTL2, BIRC5, FSTL1; five of seven canonical targets were detectable in the IBD matrix), per-sample YAP target scores were calculated in the IBD cohort. A two-sided Mann-Whitney U test for group-level comparison (IBD vs. HC). Each IBD sample’s DHR module score was determined using the mean Z-score of all 120 DHR module genes. The link between the DHR module score and the YAP target score was evaluated using Spearman rank correlation.

### Software and Reproducibility

Python 3.10; key libraries: pandas, numpy, scipy.stats, statsmodels, matplotlib, seaborn, gseapy. Analyses executed in Google Colaboratory. Code and processed results available upon request.

## Results

### IBD Mucosal Transcriptome Reveals Broad Immune Activation and Metabolic Impairment

In GSE235236, a differential expression study between IBD patients (UC + CD combined, n = 48) and healthy controls (n = 8) revealed 2,848 significantly dysregulated genes (FDR < 0.05), indicating a widespread remodelling of the mucosal transcriptome. In IBD, 1,525 were upregulated, and 1,323 were downregulated (**Figure 1A**).

**Figure 1.**
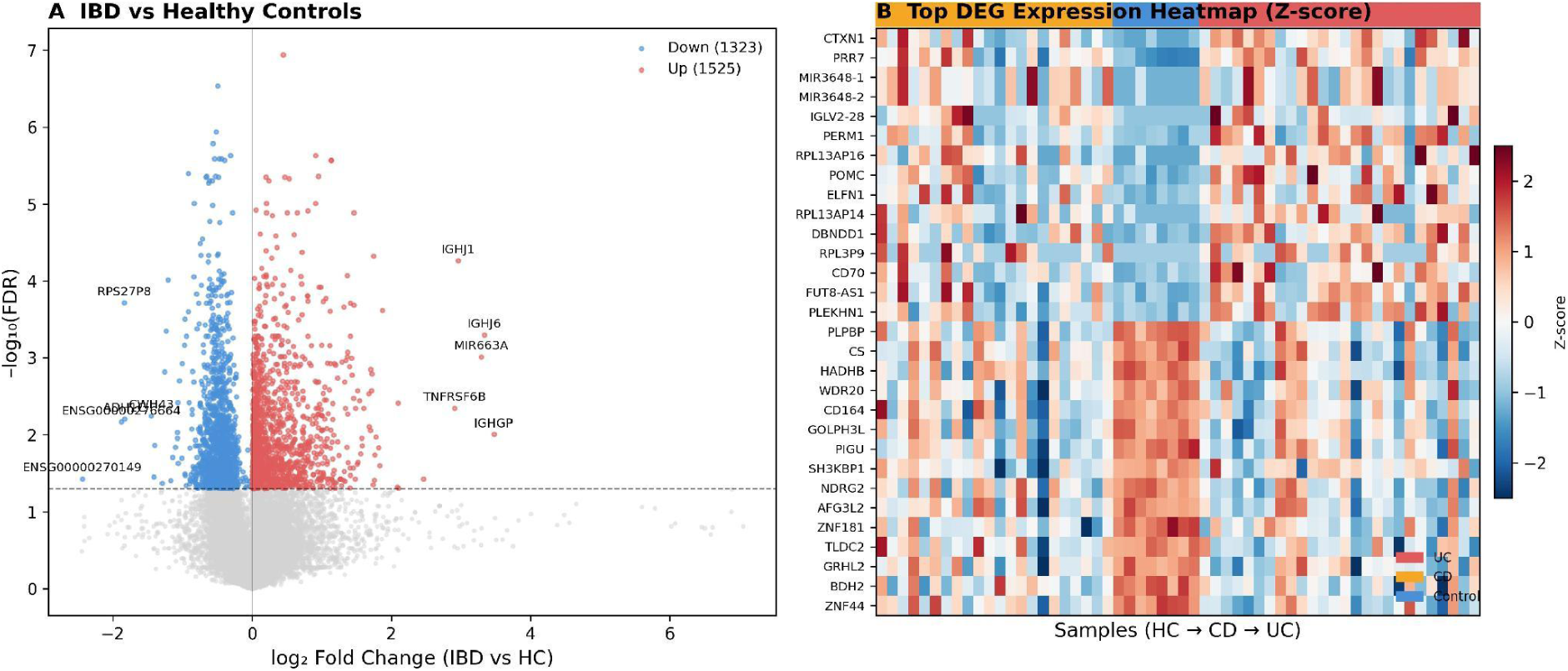
IBD mucosal differential gene expression. (A) Volcano plot: IBD (UC + CD combined, n = 48) vs. healthy controls (HC, n = 8; GSE235236). Red: upregulated in IBD (FDR < 0.05); blue: downregulated; grey: non-significant. Total significant: 2,848 (1,525 up, 1,323 down). Key genes labelled (IGHGP, IGHJ6, TNFRSF6B, ADH1C, RPS27P8, CWH43). Dashed horizontal line: FDR = 0.05. (B) Clustered heatmap of top 30 protein-coding DEGs. Row-wise Z-score normalised. Column annotation bars: group (UC: salmon; CD: orange; HC: sky blue).

Upregulated protein-coding genes prominently included immune activation and evasion transcripts. TNFRSF6B-encoding a soluble decoy receptor that neutralizes FasL and blocks apoptosis of epithelial cells-was significantly elevated (log₂FC = +2.91, FDR = 0.0045). Immunoglobulin-locus transcripts (IGHGP, log2FC = +3.48; IGHJ1, log2FC = +2.96; IGHJ6, log2FC = +3.34) reflected mucosal B-cell expansion. Alcohol dehydrogenase 1C, the main colonocyte enzyme for oxidizing microbially generated acetaldehyde, a direct mutagen, is encoded by ADH1C (log2FC = −1.83, FDR = 0.0062), one of the most downregulated genes. As explained in Section 3.4, these IBD-specific findings (TNFRSF6B and ADH1C) are notable signals of immune evasion and metabolic impairment in the inflamed mucosa; they do not meet cross-cohort concordance thresholds because they are not significantly dysregulated in TCGA Hippo-stratified tumors (TNFRSF6B: TCGA FDR = 0.054; ADH1C: TCGA FDR = 0.274) and are not members of the DHR concordant module. Additional downregulated transcripts included RPS27P8 (log2FC = −1.83) and CWH43 (log2FC = −1.45). A clustered heatmap of the top 30 protein-coding DEGs demonstrated clear IBD-control separation (**Figure 1B**).

### YAP Transcriptional Activity Elevated in IBD Mucosa

Per-sample YAP target scores (mean Z-score of detected YAP target genes: ANKRD1, AREG, AMOTL2, BIRC5, FSTL1) showed a trend toward higher values in IBD patients compared to healthy controls, though this did not reach statistical significance (Mann-Whitney U = 238.0, p = 0.293; Figure 2). The YAP target score had a median of 0.177 (IQR: −0.386 to +0.483) in IBD patients versus −0.149 (IQR: −0.817 to +0.105) in controls.

**Figure 2.**
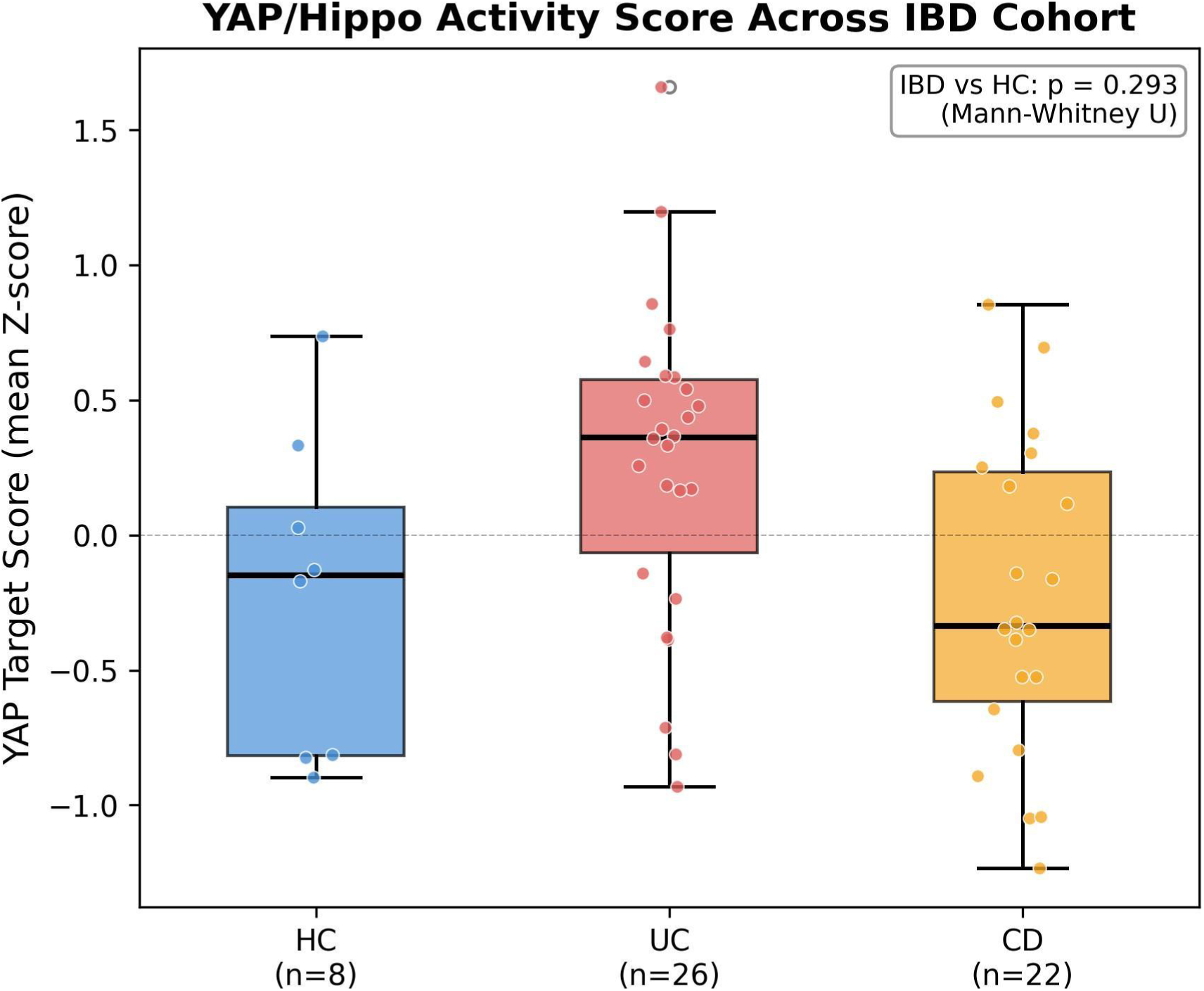
YAP transcriptional activity across the IBD cohort. Boxplot of per-sample YAP target score (mean Z-score of ANKRD1, AREG, AMOTL2, BIRC5, FSTL1) stratified by group (HC, UC, CD). IBD vs. HC: Mann-Whitney U = 238.0, p = 0.293. Individual samples overlaid. Median (IBD) = 0.177 (IQR: −0.386 to +0.483); Median (HC) = −0.149 (IQR: −0.817 to +0.105).

The small control group (n = 8) and the absence of the two most YAP-discriminatory targets (CTGF, CYR61) from the IBD expression matrix partly explain the non-significant trend. However, the highly significant proportional link between the YAP score and the whole DHR module validates the YAP program’s functional engagement across the IBD transcriptome.

### TCGA Stratification Defines Transcriptionally Distinct Hippo-High and Hippo-Low CRC Tumours

YAP target score stratification produced 148 Hippo-High (Q4) and 148 Hippo-Low (Q1) samples from 592 TCGA-CRC tumors. DEG analysis identified 10,642 significantly altered genes between the groups (FDR < 0.05; 8,061 up, 2,581 down). Hippo-Low tumors displayed comparatively higher expression of differentiation and tumor-suppressive programs, while Hippo-High tumors were characterized by enrichment for YAP/TEAD target genes and ECM remodeling transcripts; the top upregulated genes were SFRP2 (log₂FC = +4.54), COL10A1 (log₂FC = +4.33), THBS4 (log₂FC = +3.91), COMP (log₂FC = +3.89), and SFRP4 (log₂FC = +3.69) (Figure 3).

**Figure 3.**
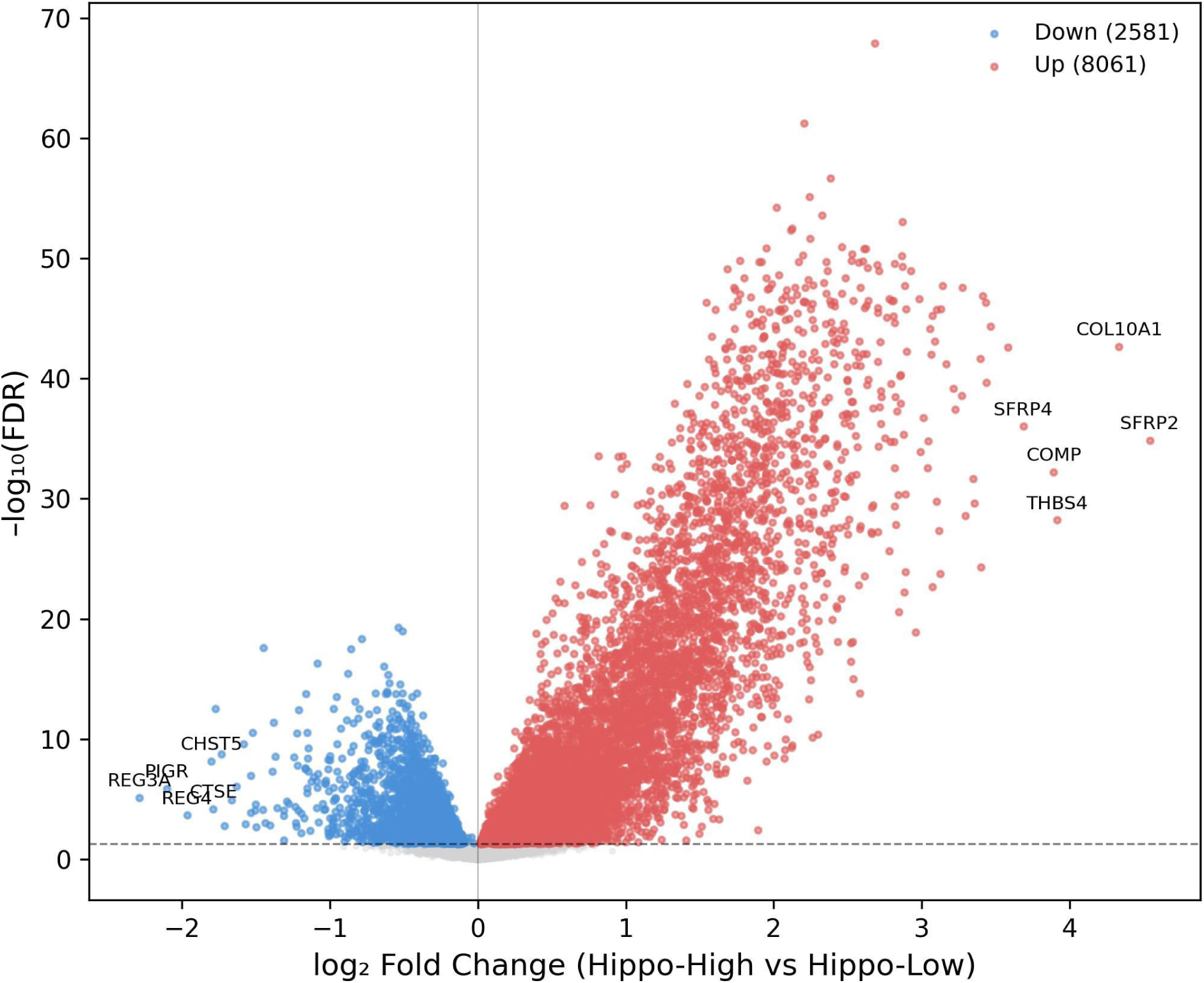
TCGA-CRC Hippo-High vs. Hippo-Low stratification. Volcano plot of DEGs between Hippo-High (Q4, n = 148) and Hippo-Low (Q1, n = 148) TCGA-CRC tumours. Total significant: 10,642 (8,061 up, 2,581 down). Top genes labelled (SFRP2, COL10A1, THBS4, COMP, SFRP4).

Importantly, the seven canonical YAP transcriptional target genes-CTGF (log₂FC = +2.21, FDR = 6.0 x 10⁻⁶²), CYR61 (log₂FC = +2.69, FDR = 1.3 x 10⁻⁶⁸), ANKRD1 (log₂FC = +1.95, FDR = 5.1 x 10⁻³²), AMOTL2 (log₂FC = +0.93, FDR = 4.4 x 10⁻³¹), and FSTL1 (log₂FC = +2.02, FDR = 6.1 x 10⁻⁵⁵)-were found to be strongly and significantly upregulated in Hippo-high CRC. These effect sizes and significance levels validate the YAP activity scoring approach. Yet, none of these canonical targets are significantly dysregulated in IBD mucosa, hence, they are not included in the concordant DHR module. This dissociation indicates that the shared IBD–CRC transcriptional program is mediated through non-canonical YAP effector genes rather than the classical CTGF/CYR61 axis.

### Cross-Cohort Overlap Defines the Dysbiosis-Hippo Response (DHR) Module

Following independent filtering, the intersection of 2,848 IBD DEGs and 10,642 TCGA DEGs yielded 147 genes significant in both cohorts, of which 120 were directionally concordant (82%), and 27 were discordant. Clustering of concordant genes in the co-upregulated and co-downregulated quadrants was validated using a log₂FC scatter plot (IBD vs. TCGA) (**Figure 4**).

**Figure 4.**
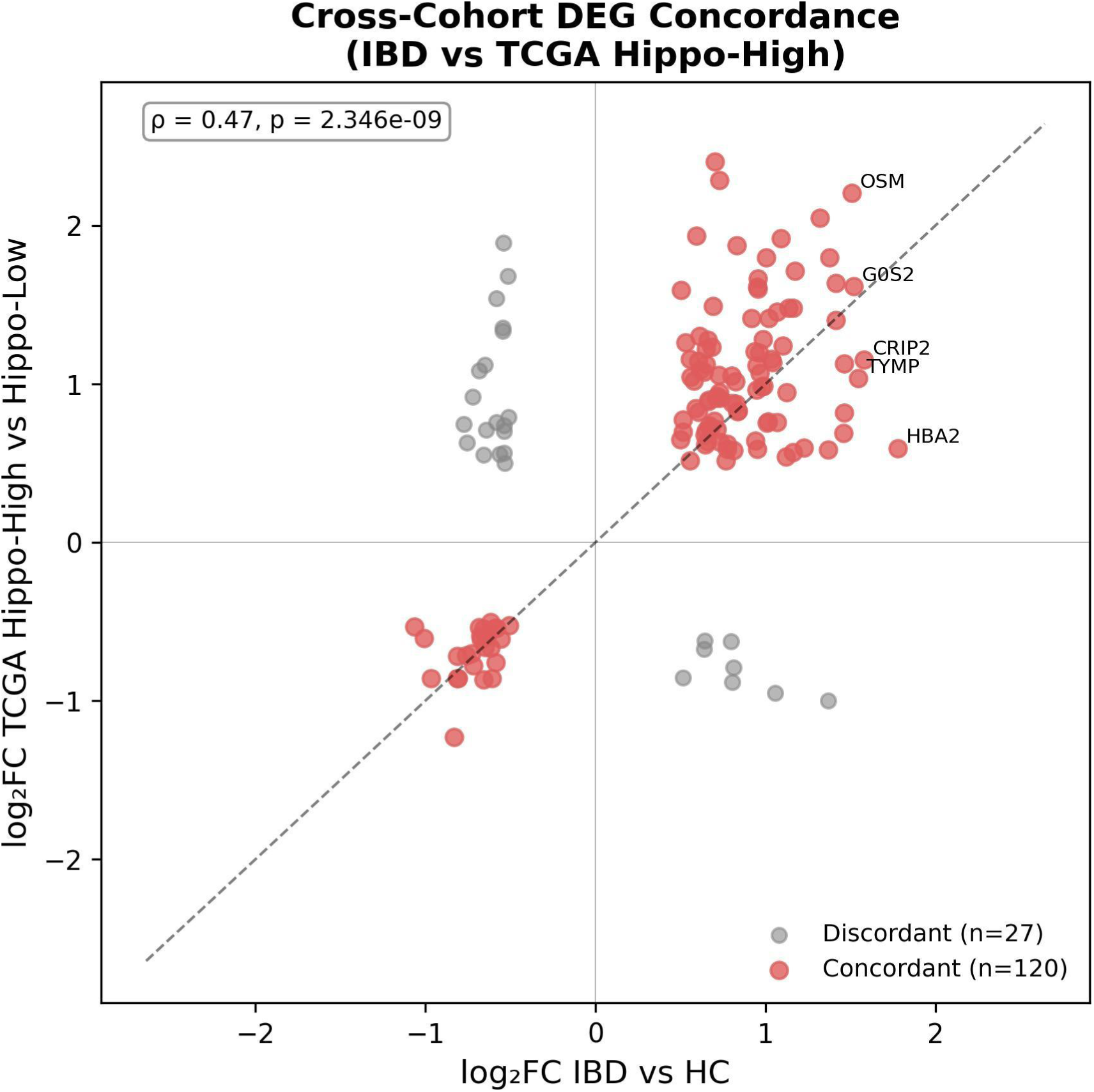
Cross-cohort log2FC concordance scatter plot. IBD log2FC (x-axis) vs. TCGA log2FC (y-axis) for all 147 genes significant in both cohorts. Red: concordant (same direction, n = 120); grey: discordant (n = 27). Diagonal reference line.

The DHR module of 120 prioritized genes was constructed by ranking these 120 concordant genes according to their cumulative S score. IBD and TCGA datasets showed identical expression patterns in heatmaps of the top 30 DHR genes (**Figure 5**). Three significant associations were identified by functional enrichment of the DHR module (MSigDB Hallmark 2020 gene sets): TNF-alpha Signaling via NF-kB (adjusted p = 0.006, combined score = 53.6, 7/200 genes), Inflammatory Response (adjusted p = 0.013, 6/200 genes), and Myogenesis (adjusted p = 0.013, 6/200 genes) (**Figure 6**). Several top-ranked DHR genes (notably TWIST1, SNAI1, and PRRX2), which are recognized members of the Hallmark Myogenesis gene set due to their function in mesenchymal programming, exhibit mesenchymal/EMT characteristics that are reflected in myogenesis enrichment. Adjusted p < 0.05 was not obtained by any other terms.

**Figure 5.**
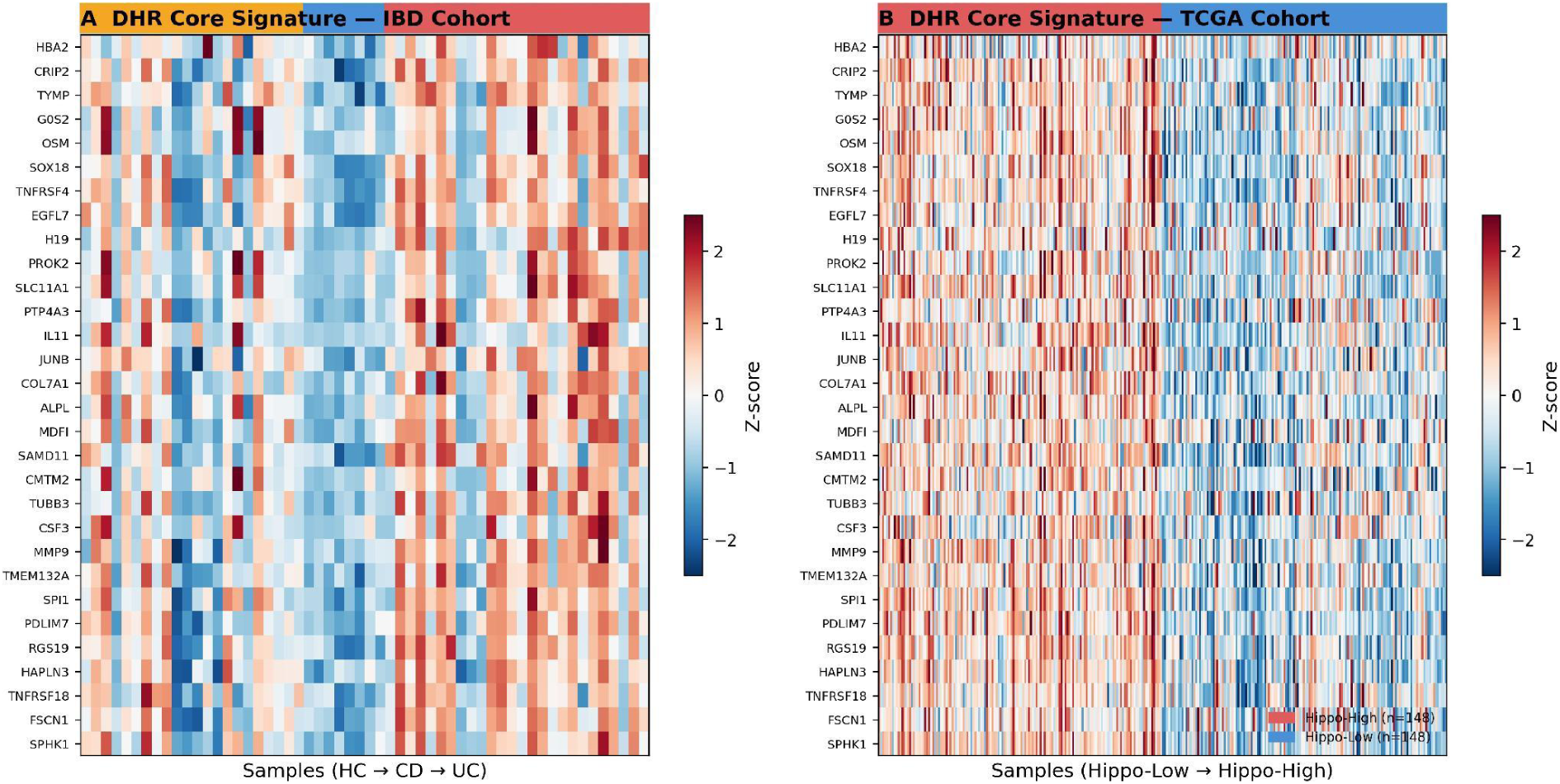
DHR module expression heatmaps. Top 30 DHR genes (ranked by S score) in (A) IBD cohort (GSE235236) and (B) TCGA-CRC cohort (Hippo stratified). Row-wise Z-score normalised.

**Figure 6.**
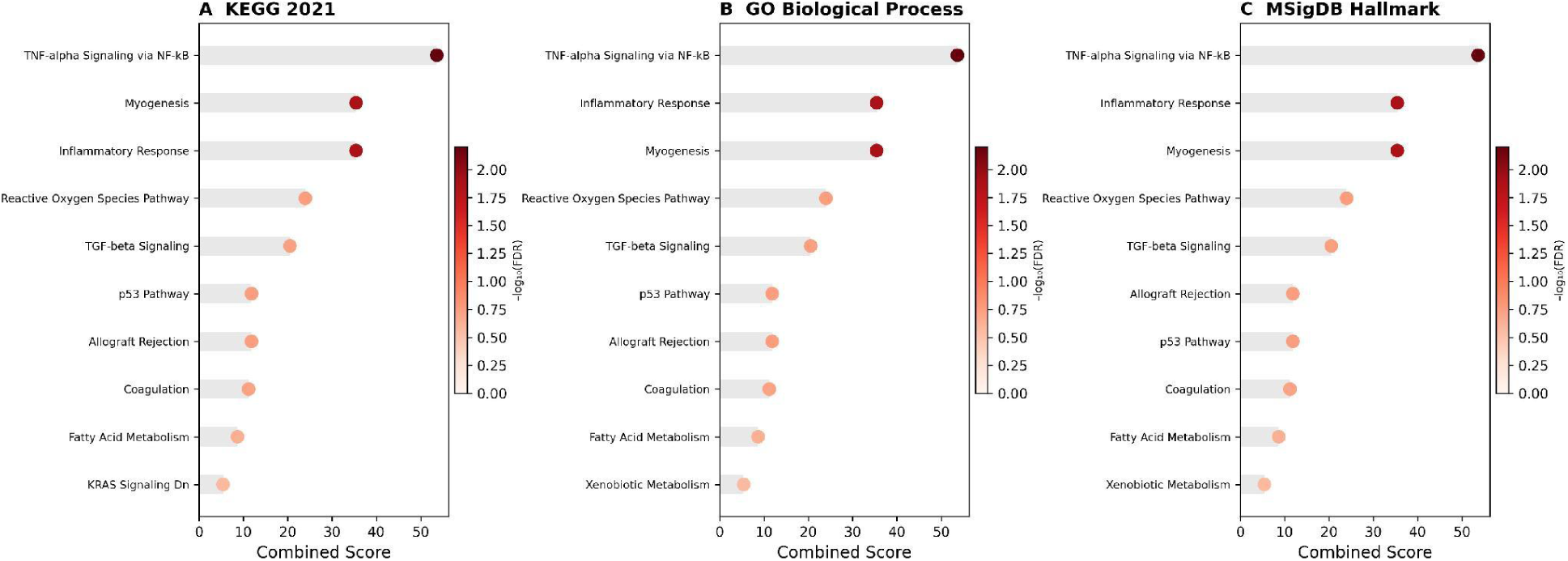
Functional enrichment of the DHR module. Dotplot of top enriched terms from MSigDB Hallmark 2020 gene sets for the 120-gene DHR module. Significant terms (adjusted p < 0.05): TNF-alpha Signalling via NF-kB (adjusted p = 0.006, 7 genes), Inflammatory Response (adjusted p = 0.013, 6 genes), and Myogenesis (adjusted p = 0.013, 6 genes). Dot size: overlap count; colour: adjusted p-value.

### DHR Module Genes Segregate Into Three Functional Archetypes Connecting Dysbiosis to YAP-Driven Oncogenesis

Three mutually reinforcing functional archetypes were identified through a systematic functional classification of the 120 concordant DHR genes, based solely on genes verified to be present in the concordant overlap (**Figure 7A**; **Table 2**).

**Figure 7.**
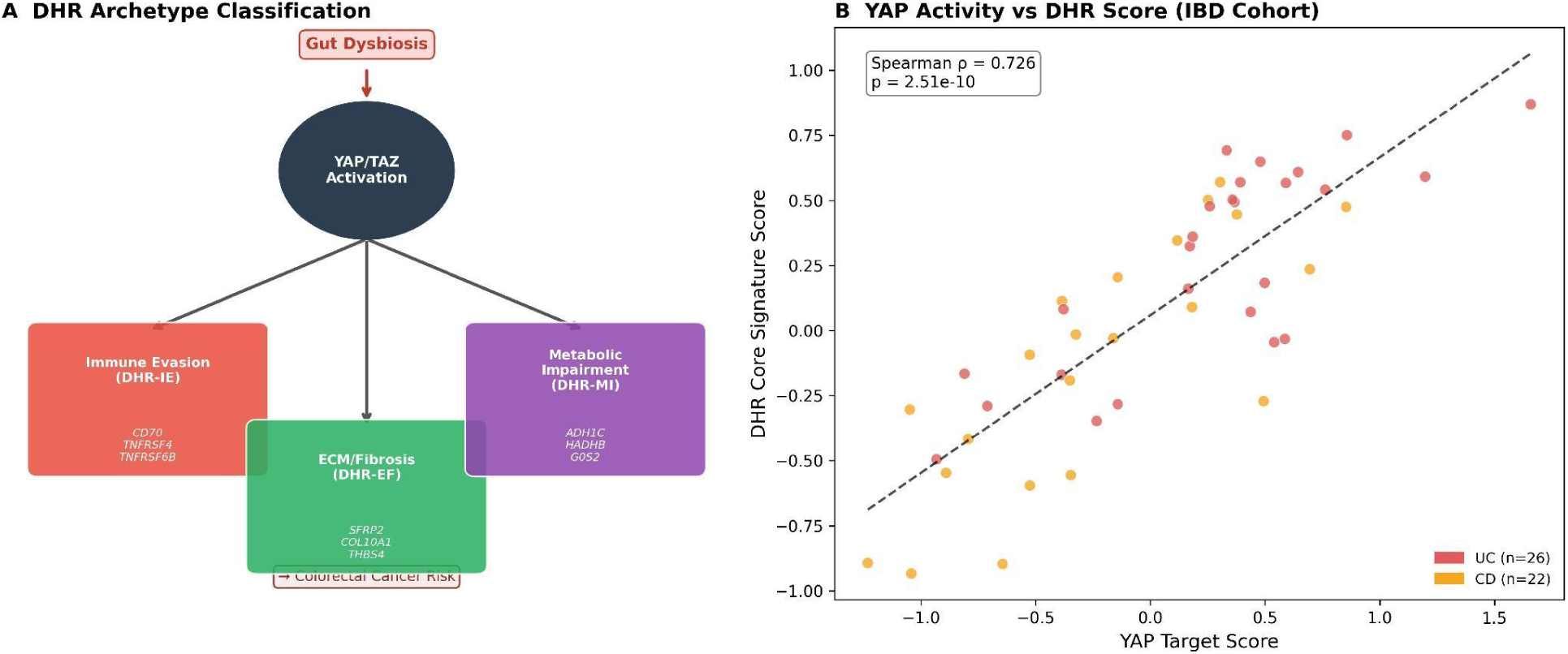
DHR gene archetypes and the YAP-DHR correlation. (A) Schematic of the three DHR functional archetypes (Innate Immune Activation, EMT/Invasive Priming, TME Immune Regulation) with arrows from gut dysbiosis through YAP activation to CRC risk. (B) Scatter plot of per-sample YAP target score vs. DHR module score across all IBD samples (n = 56; UC: red, CD: orange, HC: blue); Spearman rho = 0.726, p = 2.5 x 10-10. Dashed line: linear regression trend.

**Table 1.**
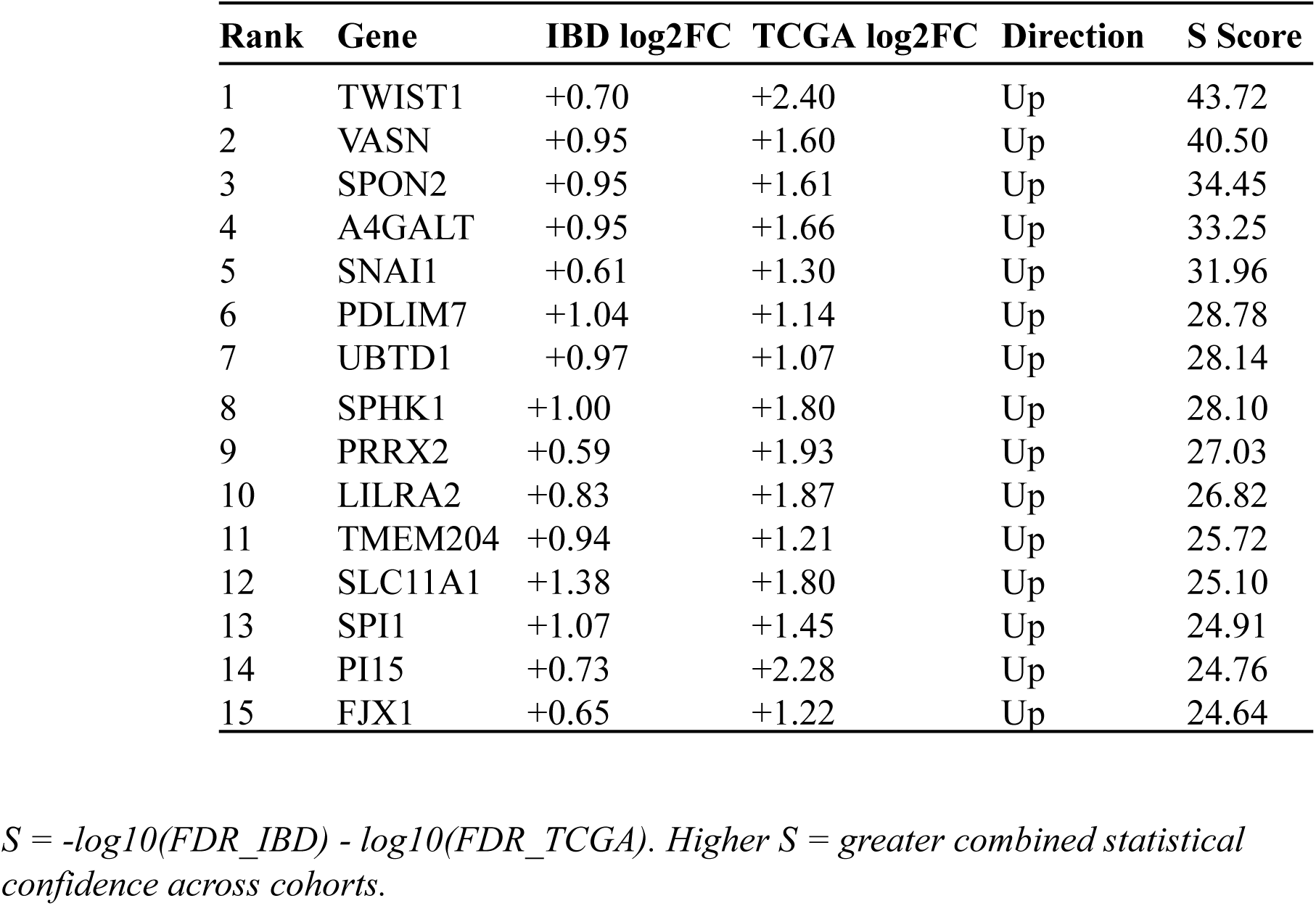
Top 15 ranked genes of the DHR module (ranked by combined S score)

**Table 2.** DHR gene functional archetype annotations (selected representative genes)

| Gene | Archetype | IBD log2FC | TCGA log2FC |
| --- | --- | --- | --- |
| SPI1 | Innate Immune Activation | 1.07 | 1.45 |
| OSM | Innate Immune Activation | 1.51 | 2.2 |
| IL11 | Innate Immune Activation | 1.32 | 2.05 |
| LILRA2 | Innate Immune Activation | 0.83 | 1.87 |
| SLC11A1 | Innate Immune Activation | 1.38 | 1.8 |
| TWIST1 | EMT / Invasive Priming | 0.7 | 2.4 |
| SNAI1 | EMT / Invasive Priming | 0.61 | 1.3 |
| PRRX2 | EMT / Invasive Priming | 0.59 | 1.93 |
| MMP9 | EMT / Invasive Priming | 1.09 | 1.92 |
| VASN | TME Immune Regulation | 0.95 | 1.6 |
| SPON2 | TME Immune Regulation | 0.95 | 1.61 |
| TNFRSF4 | TME Immune Regulation | 1.46 | 1.13 |
| PDCD1 | TME Immune Regulation | 0.67 | 0.74 |

Archetype 1: Innate Immune Activation. Genes encoding myeloid inflammatory effectors, which were all co-upregulated in both IBD mucosa and Hippo-High CRC, made up the biggest functional cluster. The major myeloid transcription factor that controls the identity of macrophages and dendritic cells is encoded by SPI1 (PU.1; IBD log2FC = +1.07, TCGA log2FC = +1.45). The IL-6-family cytokine OSM (Oncostatin M; IBD log2FC = +1.51, TCGA log2FC = +2.20) stimulates STAT3 and promotes epithelial plasticity. IL11 (IBD log₂FC = +1.32, TCGA log₂FC = +2.05) similarly signals through gp130/STAT3 and is a known cause of CRC metastasis [21]. Activated myeloid populations are identified by LILRA2 (IBD log₂FC = +0.83, TCGA log₂FC = +1.87); a macrophage iron/proton co-transporter essential to phagosomal antimicrobial killing is encoded by SLC11A1/NRAMP1 (IBD log₂FC = +1.38, TCGA log₂FC = +1.80). NCF1 and NCF1C (NADPH oxidase subunits) are co-upregulated in both cohorts, indicating that inflammatory and tumor-associated myeloid cells produce reactive oxygen species.

Archetype 2: Invasive Priming and Epithelial-Mesenchymal Transition (EMT). TWIST1, a bHLH transcription factor and master regulator of EMT that suppresses E-cadherin and activates mesenchymal gene programs, is the highest-ranked DHR gene by combined S score (S = 43.72; IBD log2FC = +0.70, TCGA log2FC = +2.40). The zinc-finger EMT transcription factor SNAI1 (Snail; IBD log2FC = +0.61, TCGA log2FC = +1.30) directly represses epithelial tight junction proteins when it is upregulated. PRRX2 (IBD log₂FC = +0.59, TCGA log₂FC = +1.93) is a paired-related homeobox factor enriched in mesenchymal stem cell programs and linked to CRC invasion. MMP9 (IBD log₂FC = +1.09, TCGA log₂FC = +1.92) encodes the gelatinase that degrades type IV collagen in basement membranes, a prerequisite for tumor cell intravasation. FSCN1 (IBD log2FC = +1.01, TCGA log2FC = +0.75) bundles filamentous actin at invadopodial tips. COL7A1 (IBD log₂FC = +1.17, TCGA log₂FC = +1.71) remodels the sub-epithelial anchoring fibril network. Field cancerization, in which pre-malignant transcriptional priming for invasion takes place before overt histological change, is consistent with the co-upregulation of this cohesive EMT program in non-dysplastic IBD mucosa and YAP-high CRC.

Archetype 3: Tumor Microenvironment Immune Regulation. A third archetype comprised secreted and cell-surface immune modulators co-elevated in both settings. VASN (Vasorin; IBD log2FC = +0.95, TCGA log2FC = +1.60) is a leucine-rich repeat protein that sequesters TGF-β, modulating epithelial–stromal crosstalk. A CRC immune microenvironment regulator, SPON2 (Mindin; IBD log₂FC = +0.95, TCGA log₂FC = +1.61), is an ECM-associated integrin ligand that stimulates macrophage adhesion [22]. co-stimulatory receptors TNFRSF4/OX40 (IBD log₂FC = +1.46, TCGA log₂FC = +1.13) and TNFRSF18/GITR (IBD log₂FC = +1.01, TCGA log2FC = +0.76) are expressed on activated T cells; their co-upregulation may reflect tumor-infiltrating lymphocyte enrichment in both contexts and has direct implications for OX40/GITR agonist immunotherapy. The co-upregulation of PDCD1/PD-1 (IBD log₂FC = +0.67, TCGA log₂FC = +0.74) in both cohorts suggests that PD-1 expression reflects a shared exhaustion-like T cell landscape linking chronic gut inflammation to established CRC. In immunologically hot tumors, CD70 (IBD log2FC = +0.80, TCGA log2FC = +1.05) encodes the CD27 ligand and stimulates NK and T-cell activation.

### YAP Target Activity Quantitatively Predicts DHR Module Expression in IBD

A highly significant positive correlation (rho = 0.726, p = 2.5 x 10-10; **Figure 7B**) was found using Spearman rank correlation between per-sample YAP target scores and DHR module scores across all IBD samples (n = 56). The degree of YAP engagement in inflammatory mucosa is quantitatively linked to activation of the entire cross-cohort oncogenic gene network, as demonstrated by the proportionally higher expression of the cancer-associated DHR program in IBD samples with higher YAP transcriptional activity.

## Discussion

Our transcriptomic data provide, to our knowledge, the first systematic transcriptomic evidence that gut dysbiosis switches on a Hippo/YAP-mediated oncogenic gene programme; the DHR module, in colonic mucosa that is still histologically normal. We primarily contribute DHR gene classification into three evidence-defined archetypes, each generating testable mechanistic predictions. Each archetype represents an independent co-occurring process: innate immune activation, EMT priming, and microenvironmental immune regulation. Canonical YAP target genes (CTGF, CYR61, ANKRD1, AMOTL2, FSTL1) are strongly induced in Hippo-High CRC yet are essentially absent from IBD mucosa) whereas the actual DHR module is built on non-canonical effectors. This demonstrates that the IBD-CRC shared transcriptome reflects upstream regulatory convergence operating through distinct, previously unrecognised gene networks.

The innate immune activation archetype pivots on SPI1/PU.1, the principal transcription factor specifying monocyte, macrophage, and dendritic cell identity. Its co-upregulation in IBD mucosa and Hippo-High CRC indicates shared expansion or polarisation of myeloid populations in both diseases. That interpretation is reinforced by the co-elevation of downstream SPI1 targets: LILRA2 (inhibitory Ig-like receptor modulating myeloid activation thresholds), SLC11A1/NRAMP1 (divalent metal transporter in macrophage phagosomes), and NCF1/NCF1C (p47-phox and its centrosomal paralogue, core components of the NADPH oxidase superoxide-generating complex). We also found OSM and IL11 co-elevated in both cohorts-tellingly, gp130-signalling IL-6 family cytokines. This matters because OSM activates STAT3 in colonocytes and has been shown to disrupt tight junction integrity and induce EMT-like plasticity [21], while IL11 drives CRC peritoneal metastasis through STAT3 [21]. The convergence of myeloid-derived cytokine signalling in IBD and YAP-active CRC therefore implies a conserved innate immune-to-epithelial axis that may progressively condition the epithelium toward oncogenic transformation.

TWIST1 heads the EMT archetype as the top-ranked DHR gene. Its upregulation in non-dysplastic IBD mucosa is remarkable. TWIST1 represses E-cadherin by binding E-box elements in its promoter, activates N-cadherin, vimentin, and fibronectin, and confers resistance to anoikis, together constituting the invasive cell phenotype [23]. Its co-upregulation in Hippo-high CRC (log₂FC = +2.40) indicates that the programme engaged in inflamed epithelium is the same one deployed at high amplitude in established tumors with YAP activity. SNAI1/Snail reinforces this by repressing the tight junction proteins claudin and occludin, directly disrupting the epithelial barrier. PRRX2 adds a mesenchymal stem-like dimension, as it marks the most invasive subpopulation of CRC cells [24].

MMP9 (gelatinase B), which cleaves type IV collagen in basement membranes and is a known effector of macrophage-driven tumor invasion in colorectal cancer, is present in this archetype and links EMT transcription to ECM degradation. The field cancerization theory is supported by the co-upregulation of several EMT genes in IBD and Hippo-High CRC [17]: the inflamed mucosal field is transcriptionally pre-conditioned for invasion before any histological change is detectable. DHR module EMT gene scores from colonoscopic biopsies may serve as molecular risk stratifiers independent of dysplasia, with direct clinical implications.

Immune checkpoint and co-stimulatory molecules are strikingly co-upregulated in both IBD mucosa and Hippo-high CRC, according to the third archetype. PDCD1/PD-1 expression is elevated in both conditions, indicating a shared T-cell exhaustion-like state, consistent with the established efficacy of PD-1 blockade in microsatellite-instability-high CRC and with growing evidence of T-cell dysfunction in chronic IBD. Clinically actionable is the co-elevation of TNFRSF4/OX40 and TNFRSF18/GITR, agonist targets in clinical cancer immunotherapy trials, with PD-1 in both cohorts. This suggests a shared co-inhibitory/co-stimulatory immune circuit in gut inflammation and YAP-active CRC that may be targetable with combination immunotherapy.

VASN (vasorin) modifies the stromal signaling environment by sequestering TGF-β via its leucine-rich repeat domain. SPON2 (Mindin) is an ECM-associated opsonin that stimulates macrophage adhesion, phagocytosis, and M1-like polarization [22]. Recent research has identified SPON2 as a microenvironmental determinant of CRC immune infiltration, and its co-upregulation in both situations may signify shared recruitment of classically activated macrophages as part of the innate inflammatory response. Reading together, this archetype describes an immune regulatory program that represents both YAP-active tumor immune microenvironment organization (CRC) and chronic inflammation (IBD).

The lack of classical YAP transcriptional targets in the DHR-concordant module is a significant finding of our investigation. CTGF, CYR61, ANKRD1, AMOTL2, and FSTL1 are among the most statistically significant DEGs in Hippo-High vs. Hippo-Low TCGA tumors, with FDRs ranging from 10-31 to 10-68, confirming robust canonical TEAD target activation in established CRC. Yet, there is not a clear dysregulation of any of these genes in the mucosa of IBD. This dissociation has biological significance because it implies that canonical YAP/TEAD target induction is a transcriptional response specific to the tumor stage that relies on oncogenic co-factors (such as TEAD amplification, RAS mutation-driven YAP nuclear exclusion reversal, and chromatin remodeling) that are not present in the pre-malignant inflammatory epithelium. Instead, independent of classical CTGF/CYR61 signaling, the DHR module captures the upstream-convergent transcriptional state-innate immune activation, EMT priming, and checkpoint engagement-that both IBD and YAP-active CRC share. This non-canonical program constitutes a mechanistically distinct, uncharacterized molecular bridge between dysbiosis and malignancy.

The DHR module, a gene program expressed in non-dysplastic IBD mucosa whose expression pattern resembles YAP-active CRC, is consistent with the field carcinogenesis framework [17]. A computationally calculated YAP activity score from a colonoscopic biopsy could function as a continuous risk predictor for future colorectal cancer (CRC) without requiring any histological abnormality, according to the quantitative association between YAP target score and DHR module score (rho = 0.726, p = 2.5 x 10-10).

A few limitations are worth acknowledging. First, the small HC group (n = 8) limits statistical power; the non-significant YAP score difference (p = 0.293) should be interpreted in this context. Second, the TCGA cohort lacks matched adjacent normal tissue. Third, DHR gene archetype annotations assigned to DHR genes will require experimental validation. Fourth, causality cannot be established by transcriptome correlations. Fifth, our overlap approach does not detect genes that have large effects in only one of the two cohorts.

### Future Directions

Every archetype raises a different research question. For Archetype 1, would genetic or pharmacological inhibition of SPI1/PU.1 or STAT3 (downstream of OSM/IL11) reduce CRC incidence or DHR module expression in murine colitis models? For Archetype 2: can TWIST1 or SNAI1 inhibitors suppress field cancerization gene programs in IBD organoids? For Archetype 3: do OX40/GITR agonists combined with PD-1 blockade show synergistic activity in IBD-associated CRC models? Longitudinal IBD cohorts should also be examined for whether the DHR module score predicts subsequent dysplasia or CRC, and single-cell RNA-seq will identify which specific epithelial and stromal cell populations express each archetype. Future research is necessary to determine if the IBD-specific DEGs TNFRSF6B (log₂FC = +2.91) and ADH1C (log₂FC = −1.83) represent CRC-subtype-specific or microenvironment-contextual effects and why they are not concordant in TCGA Hippo-stratified tumors.

## Supplementary Materials

**Supplementary Table S1.** Full IBD DEG list (2,848 significant genes). File: IBD_DEG_IBD_vs_Control.csv.

**Supplementary Table S2.** Full TCGA Hippo-High vs. Hippo-Low DEG list (10,642 significant genes). File: TCGA_DEG_HippoHigh_vs_HippoLow.csv.

**Supplementary Table S3.** Complete ranked DHR module gene list (120 genes). File: core_signature_genes.csv.

**Supplementary Table S4.** Full functional enrichment results. File: core_signature_enrichment.csv.

**Supplementary Figure S1.** Distribution of YAP target scores across TCGA-CRC tumours (n = 592), showing Q1/Q4 stratification thresholds.

**Supplementary Figure S2.** Volcano plots of UC vs. HC and CD vs. HC individual subgroup analyses.

## Data and Code Availability

The accession numbers of used publicly available datasets are mentioned in Methods section and analysis based notebooks are provided at https://github.com/GeneticCodon/Colorectal_Hippo_Dysbiosis

## Ethics Approval Statement

Not applicable. This study analyzed publicly available, de-identified datasets. The data were retrieved from NCBI GEO and accession ID is mentioned in the Methodology section.

## Permissions for Reproducibility

All permissions are granted to reproduce material from other sources

